# A Brief Report on Coccidioidomycosis and Malignancies in an Endemic Area

**DOI:** 10.1101/2025.07.05.25330009

**Authors:** Rupam Sharma, Kevin T. Dao, Aishwarya Saripalli, Akriti Chaudhry, Royce H. Johnson, Carlos D’Assumpcao, Arash Heidari, Everardo Cobos, Leila Moosavi

## Abstract

Coccidioidomycosis, a fungal infection caused by *coccidioides* species, is endemic to regions with arid climates, such as the southwestern United States. This disease predominantly affects the respiratory system by may disseminate in immunocompromised individuals. Cancer patients, particularly those undergoing immunosuppressive therapies, are at heightened risk for opportunistic infections including coccidioidomycosis. This paper explores the complex interplay between coccidioidomycosis and cancer in an endemic area and the impact of co-occurrence on patient outcomes.

## Introduction

*Coccidioides* species are thermal dimorphic soil fungal pathogens irregularly distributed in much of the southwestern United States adjacent Mexico and similar but restricted areas of the rest of Latin America. There is substantial concern that the distribution of *Coccidioides* is expanding, potentially increasing the threat of disease in an increased population. There is evidence that immunocompromising illness and/or therapeutic interventions increase the frequency and severity of *Coccidioides* illness.

Coccidioidal infection is almost always acquired by inhalation of airborne arthroconidia (spores) (3). 60% of infections are asymptomatic (3). The majority of symptomatic illness is respiratory. Three out of four symptomatic cases are not diagnosed. The diagnosed cases tend to be more the severe and persistent pneumonic disease. A small percentage of infections (1%) develop as disseminated disease (4). These cases may present as denovo infectious process anywhere in the body. This may occur simultaneously with or without a pneumonic antecedent.

Malignant disease is a global health problem. Despite major advances in prevention, diagnosis and treatment it remains a major cause of death. Others have published on epidemiologic risk factors on the risk of developing symptomatic, severe pulmonary and disseminated coccidioidomycosis in various populations (2,4). These and others have observed that immunocompromising illness including cancer is associated with more frequent severe and disseminated disease (2).

The purpose of this paper is to explore the nature of the disease relationship of cancer and cocci in an academic public hospital serving a substantial population with underlying problematic determinants of health.

The aim of this study is to evaluate the risk to patients who are diagnosed with coccidiomycosis before and after cancer.

## Methods

This study was approved by the institutional review board, Kern Medical. A literature review was undertaken in PubMed and Google Scholar. Search terms included coccidioidomycosis, coccidioidomycosis and cancer. Patients were abstracted from the medical records of Kern Medical from Jan 2016 – March 2022 using ICD 10 codes. 3342 patients were diagnosed with malignant disease. 1961 patients were diagnosed with Coccidioidomycosis. Inclusion criteria were a diagnosis of diagnosed malignancy and either antecedent or post malignancy coccidioidomycosis. Malignancy was diagnosed based on histopathology or other agreed upon testing. The diagnosis of coccidioidomycosis was based on a compatible clinical illness, plus IgG antibody by immunodiffusion or complement fixation, and/or histopathology with endosporulating spherules from Kern County Public Health Department or University of California, Davis.

Exclusion criteria included absence of inclusion criteria, age under 18, pregnancy, human immunodeficiency virus positive patients, a comorbid immunocompromising illness (other than diabetes mellitus), tuberculosis or incarcerated individuals. 53 patients were analyzed. Patient’s charts were abstracted for: age, race, sex, comorbidities, cancer diagnosis date, chemotherapy, radiation therapy, immunotherapy, hormonal therapy, coccidioidomycosis date of diagnosis, coccidioidomycosis disease category, whether the coccidioidomycosis was antecedent, or post malignancy and pulmonary vs extrapulmonary disseminated coccidiomycosis.

## Results

The demographics of the 53 patients are shown in Table 1. We found 32 male patients, 21 female patients, approximately one half the patients were Latinx. This is in keeping with the general population of Kern County and the people we serve. Table 3 describes the cancer types which included prostate cancer, renal cell carcinoma, basal and squamous cell carcinoma of skin, thyroid cancers, gynecological malignancies, melanomas, genitourinary malignancies and gastroenterology malignancies (including esophageal and colorectal). Tables 4 and 5 showed 27 patients with cancer diagnosis antecedent to Coccidiomycosis diagnosis, 26 of whom had pulmonary coccidiomycosis, and one had disseminated Coccidiomycosis. 26 patients had cancer diagnosis after coccidiomycosis diagnosis, 13 of whom had pulmonary coccidiomycosis and 13 with disseminated coccidiomycosis. We had a large number of patients with diabetes and they were equally distributed in those who had coccidioidomycosis pre and post malignancy.

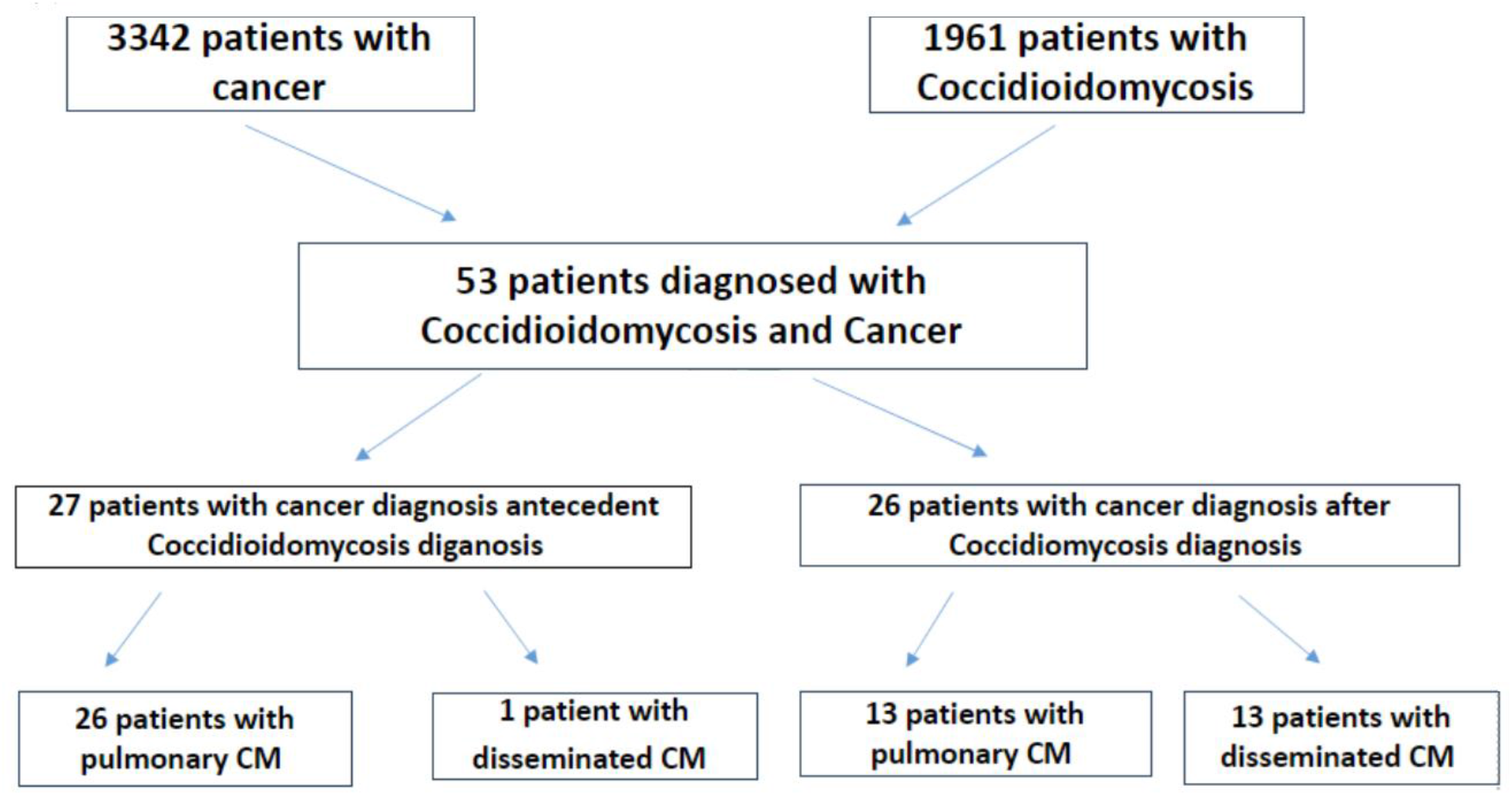

**Table 1:** Demographic Characteristics of 53 Patients with Malignancies and Coccidioidomycosis.

|  | Patients No. |  |
| --- | --- | --- |
|  | No. | % |
| **Gender** |  |  |
| Female | 21 | 39.6 % |
| Male | 32 | 60.3 % |
| **Ethnicity** |  |  |
| Hispanic | 27 | 50.9 % |
| White | 18 | 33.9 % |
| African American | 5 | 9.4 % |
| Asian/Filipino | 3 | 5.6 % |
| **Age of Cancer** |  |  |
| 20-29 | 1 | 1.8 % |
| 30-39 | 4 | 7.5 % |
| 40-49 | 11 | 20.7 % |
| 50-59 | 22 | 41.5 % |
| 60-69 | 10 | 18.8 % |
| >70 | 5 | 9.4 % |
| **Age of Cocci Dx** |  |  |
| 20-29 | 3 | 5.6 % |
| 30-39 | 6 | 11.3 % |
| 40-49 | 14 | 26.4 % |
| 50-59 | 14 | 26.4 % |
| 60-69 | 11 | 20.7 % |
| >70 | 5 | 9.4 % |

**Table 2:** Coccidioidomycosis Characteristics of 53 Patients with Cancer.

|  | Patients No. |  |
| --- | --- | --- |
|  | No. | % |
| **Location** |  |  |
| Pulmonary | 39 | 74 |
| Disseminated | 14 | 26 |
| **Treatment** | **Treatment** | **Treatment** |
| Fluconazole | 43 |  |
| Isavuconazonium | 6 |  |
| Itraconazole | 5 |  |
| Voriconazole | 5 |  |
| Posaconazole | 2 |  |

**Table 3:** Cancer Characteristics of 53 Patients with Coccidioidomycosis.

| Cancer | Number of Patients | Treatment |  |  | Coccidioidomycosis |  |
| --- | --- | --- | --- | --- | --- | --- |
|  |  | Surgery | Chemo | Radiation | Pulmonary | Disseminated |
| Prostate Cancer | 14 | 8 | 2 | 5 | 10 | 5 |
| Renal Carcinoma | 7 | 7 |  |  | 4 | 3 |
| Basal Cell Carcinoma | 2 | 2 |  |  | 2 |  |
| Thyroid | 4 | 4 |  | 1 | 3 | 1 |
| Gynecological | 6 | 5 | 3 | 3 | 6 |  |
| Melanoma | 3 | 3 |  |  | 2 | 1 |
| GI | 7 | 5 | 4 | 1 | 6 | 1 |
| Squamous cell carcinoma | 4 | 4 |  |  | 1 | 3 |
| Metastatic neuroendocrine | 1 | 1 | 1 |  | 1 |  |
| Mantle cell lymphoma | 1 |  | 1 |  | 1 |  |
| Microcystic adnexal carcinoma | 1 | 1 |  |  |  | 1 |
| Leiomyosarcoma | 1 | 1 |  |  | 1 |  |
| Pheochromocytoma | 1 | 1 |  |  |  | 1 |
| Bladder | 1 | 1 |  |  |  | 1 |
| Total | 53 | 43 | 11 | 10 | 37 | 17 |

**Table 4:**
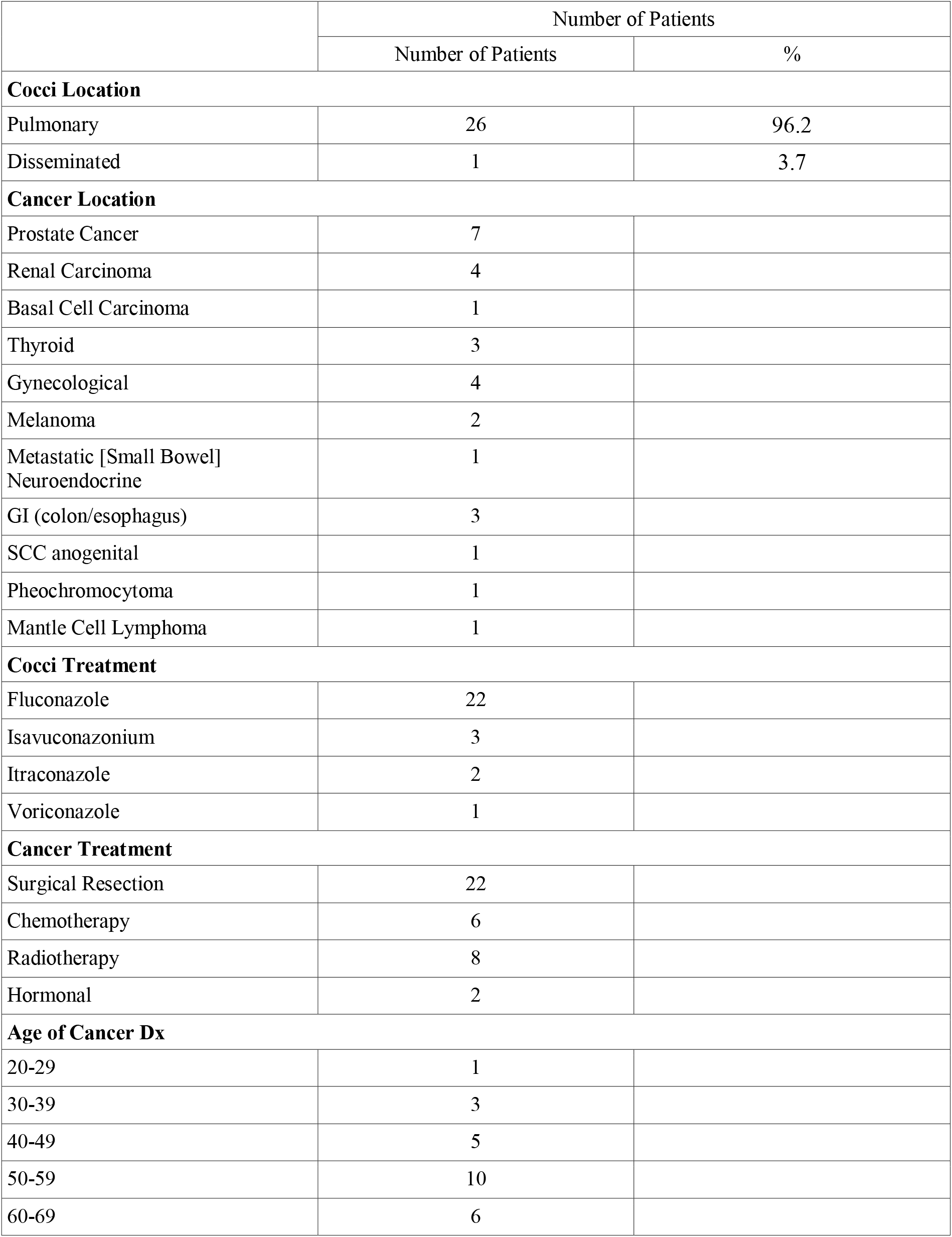
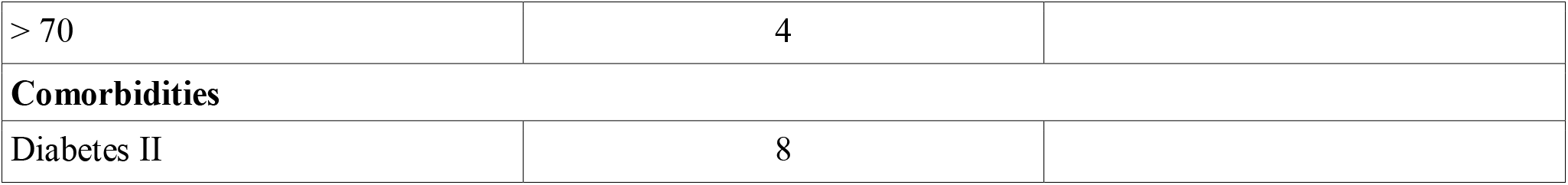
Characteristics of 27 Patients with Cancer Prior Coccidioidomycosis.

|  | Number of Patients |  |
| --- | --- | --- |
|  | Number of Patients | % |
| **Cocci Location** |  |  |
| Pulmonary | 26 | 96.2 |
| Disseminated | 1 | 3.7 |
| **Cancer Location** |  |  |
| Prostate Cancer | 7 |  |
| Renal Carcinoma | 4 |  |
| Basal Cell Carcinoma | 1 |  |
| Thyroid | 3 |  |
| Gynecological | 4 |  |
| Melanoma | 2 |  |
| Metastatic [Small Bowel]<br>Neuroendocrine | 1 |  |
| GI (colon/esophagus) | 3 |  |
| SCC anogenital | 1 |  |
| Pheochromocytoma | 1 |  |
| Mantle Cell Lymphoma | 1 |  |
| **Cocci Treatment** |  |  |
| Fluconazole | 22 |  |
| Isavuconazonium | 3 |  |
| Itraconazole | 2 |  |
| Voriconazole | 1 |  |
| **Cancer Treatment** |  |  |
| Surgical Resection | 22 |  |
| Chemotherapy | 6 |  |
| Radiotherapy | 8 |  |
| Hormonal | 2 |  |
| **Age of Cancer Dx** |  |  |
| 20-29 | 1 |  |
| 30-39 | 3 |  |
| 40-49 | 5 |  |
| 50-59 | 10 |  |
| 60-69 | 6 |  |
| > 70 | 4 |  |
| **Comorbidities** |  |  |
| Diabetes II | 8 |  |

**Table 5:** Characteristics of 26 Patients with Coccidioidomycosis Prior Cancer.

|  | Number of Patients |  |
| --- | --- | --- |
|  | Number of Patients | % |
| **Cocci Location** |  |  |
| Pulmonary | 13 | 50 |
| Disseminated | 13 | 50 |
| **Cancer Location** |  |  |
| Renal Cell Carcinoma | 3 |  |
| Prostate Cancer | 8 |  |
| Gynecological | 2 |  |
| GI Cancer | 4 |  |
| Melanoma | 1 |  |
| Squamous Cell Carcinoma | 3 |  |
| Basal Cell Carcinoma | 1 |  |
| Bladder Cancer | 1 |  |
| Leiomyosarcoma | 1 |  |
| Microcystic adnexal carcinoma | 1 |  |
| Thyroid | 1 |  |
| **Cocci Treatment** |  |  |
| Fluconazole | 21 |  |
| Isavuconazonium | 3 |  |
| Itraconazole | 3 |  |
| Voriconazole | 4 |  |
| Posaconazole | 2 |  |
| **Cancer Treatment** |  |  |
| Surgical Resection | 20 |  |
| Chemotherapy | 5 |  |
| Radiotherapy | 2 |  |
| Hormonal | 1 |  |
| **Age of Cancer Dx** |  |  |
| 20-29 | 2 |  |
| 30-39 | 4 |  |
| 40-49 | 9 |  |
| 50-59 | 5 |  |
| 60-69 | 5 |  |
| > 70 | 1 |  |
| **Comorbidities** |  |  |
| Diabetes II | 11 |  |

## Discussion

This study was aimed to evaluate the interface of two common diseases at an academic public hospital. The data which are limited due to sample size suggest that there is risk of coccidioidomycosis in persons who develop malignancy. That risk appears to be small in persons with antecedent coccidioidomycosis but substantial in persons who develop clinical coccidioidomycosis post malignancy. However, the absolute risk to any given person appears to be small.

This study differs from the Fitterer study as they found no relationship between malignancy and coccidiomycosis infection (2). The difference is not expanded by the coincidence of diabetes mellitus. Other variables are the type of malignancies found, different (age/sex/race) and social determinants of health. Clearly, larger prospective multicenter studies of coccidioidomycosis in immunocompromised patients would advantage our understanding of the interaction between malignancy and coccidiomycosis.

The limitations of this study include social determinants, sample size, broader scope of but different malignancies than were found in the study by Blair *et al*. (8) The strength of this study is that we studied a broad sweep of malignancies. Additionally, our population has challenges with social determinants of health which may mean that this population is different than the population seen in study by Blair *et al*. Previously published study, Coccidioidomycosis in patients with hematologic malignancies only included patients with non-Hodgkin lymphoma, chronic lymphocytic leukemia and multiple myeloma which were not heavily represented in our study. Additionally, our study included a much broader scope of malignancies.

## Data Availability

All data produced in the present work are contained in the manuscript

## Acknowledgments

None.

## Ethics Approval

Ethical approval to conduct this review was obtained from the Kern Medical Institutional Review Board. IRB: 20038

